# Predictors of Postoperative pharyngeal complaints in children undergoing surgery under general anesthesia at Hawassa university comprehensive specialized hospital, a *prospective observational study*

**DOI:** 10.1101/2024.03.31.24305142

**Authors:** Adanech Shifarew Legasse, Addisu Mossie, Aschalew Besha, Nesra Mohammed

**Affiliations:** Department of Anesthesia, Faculty of Medicine, College of medicine and health science, Hawassa University, Hawassa, Sidama, Ethiopia; Department of Anatomy, Adama hospital medical college, Adama, Oromia, Ethiopia

**Keywords:** postoperative hoarseness, sore throat, hoarseness, dysphagia, postoperative throat pain, pharyngeal morbidity, pharyngeal complaints

## Abstract

**Background:** Postoperative pharyngeal complaints are common but minor complications following surgery and are commonly reported during postoperative visits. These complaints includes sore throat, dysphagia, and hoarseness, which can significantly impact a child’s comfort, overall well-being and satisfaction after surgery. Identifying and understanding the predictors of these postoperative pharyngeal complaints is crucial to improve the overall outcomes in children undergoing surgery. This study aimed to assess the predictors of postoperative pharyngeal complaints in children undergoing surgery under general anesthesia.

**Methods:** A prospective observational study was conducted among children aged 6 to 16 years who underwent emergency and elective surgical procedures under general anesthesia. The data was entered and analyzed using SPSS version 26 software packages. Univariate and multivariate analyses were performed to investigate the independent predictors of postoperative pharyngeal complaint. The postoperative pharyngeal complaints (Sore throat, dysphagia and hoarseness) were assessed at 2nd, 6th, 12th, and 24th hours postoperatively.

**Result:** A total of 102 patients were included in this study, of which 68 of them male. The result of this study showed the overall incidence of postoperative pharyngeal complaints was 32.4%; whereas the incidence of postoperative sore throat 26.5%, cough 5.9%, Postoperative hoarseness 2.9% and dysphagia 4.9% of participants. Endotracheal intubation was identified as the only independent predictors of postoperative pharyngeal complaints with p-values of 0.01 (AOR-3.846, 95% CI [1.385-10.682]).

**Conclusion:** This study revealed the overall incidence of postoperative pharyngeal complaints was 32.4%. Endotracheal intubation was identified as the only independent predictors of postoperative pharyngeal complaints in children in this study.

## INTRODUCTION

### Background

Postoperative pharyngeal complaints are common but minor complications following surgery and are commonly reported during postoperative visits. These complaints can vary in severity, ranging from mild throat irritation to severe symptoms such as incapacitating pain (sore throat), difficulty swallowing (dysphagia), and temporary voice alterations (hoarseness) (1).

General anesthesia is widely used anesthetic technique during surgical procedures in pediatric surgery to ensure the patient’s safety and comfort. However, the administration of general anesthesia has its own consequences, including alterations in upper airway physiology and anatomy which may contribute to the development of postoperative pharyngeal complaints (2 - 6).

Several risk factors of postoperative pharyngeal complaints in children undergoing surgery under general anesthesia has been proposed. These include extensive surgeries, such as tonsillectomies and craniofacial surgeries that often involve manipulation of the upper airway, leads to a higher risk of postoperative pharyngeal complaints (5). Additionally, longer surgical durations and increased intraoperative airway instrumentation have been associated with a higher incidence of these complaints (3, 6).

Children are particularly prone to experiencing postoperative sore throat (POST) and postoperative hoarseness (PH), with incidence rates ranging from 3.3% to 50.7% and 4% to 42%, respectively, in developed countries (7 - 7). Younger children may have smaller airways and less-developed airway protective mechanisms, making them more susceptible to airway irritation and injury (10). Other factors that related to the anesthesia management and technique may also contribute to the development of postoperative pharyngeal complaints. The use of endotracheal tubes (ETT) or Laryngeal mask airway (LMA) during surgery can cause direct trauma to the pharyngeal tissues, potentially leading to postoperative pharyngeal symptoms (6).

Many cases of postoperative pharyngeal complaints resolve spontaneously. However, moderate to severe cases of complaint like sore throat can be managed with a gargle containing local anesthetics, dexamethasone, and analgesics, such as benzydamine hydrochloride (8, 11-13).

Identifying and understanding the Predictors of postoperative pharyngeal complaint among pediatrics surgical patients undergoing surgery under general anesthesia helps healthcare providers and professionals to optimize anesthetic management strategies, to select appropriate airway instruments, and to develop postoperative care guidelines that aim to minimize postoperative pharyngeal complaint. The aims of this study is to assess predictors of postoperative pharyngeal complaints in children undergoing surgery under general anesthesia in resource constraint area.

## Materials and Methods

A prospective observational study was conducted at Hawassa University Comprehensive and Specialized Hospital, located 273 kilometers south of Addis Ababa, from February 2022 to June 2022 (16). This study included pediatric surgical patients with ASA class I and class II physical statuses, aged 6 to 16 years, who underwent both elective and emergency surgeries under general anesthesia during the study period. Patients with a history of upper respiratory tract infection, surgeries involving airway packing, children with learning difficulties, and children with congenital airway anomalies were excluded from the study.

### Sample size and Sampling procedures

The sample size for this study was calculated using the single population proportion formula. Assuming a proportion of 50%, a 95% confidence interval, and a margin of error of 0.05, the initial sample size was calculated to be n=384. However, based on the results of a situational analysis indicating that only 123 cases were performed within three consecutive months, it was estimated that the population size was less than 10,000.

Therefore, a correction formula was applied, and a 10% non-response rate was added. Consequently, the final sample size was determined to be n=102 pediatric surgical patients. The study participants were selected at Hawassa University Comprehensive and Specialized Hospital (HUCSH) using a consecutive sampling method, whereby all pediatric surgical patients who underwent surgery and met the inclusion criteria were included in the study.

### Ethics approval and consent to participate

The advantage and risk of the study was explained written informed consent was obtained from each study participants (caregiver or family). Support letter and ethical clearance with reference number duchm/irb/055/2022 were secured from Dilla University institutional review board

### Data Collection procedure

The data collection procedure involved the use of a structured questionnaire administered to families and children who underwent surgery under general anesthesia. Prior to the surgery, both the participants’ families and the children themselves were provided with a detailed explanation of the study. Information about the study’s advantages, disadvantages, and objectives was provided to the parents and participants in English and translated into Amharic. Two data collectors and one supervisor were involved in the data collection process.

Preoperatively, written informed consent was obtained from the participants’ families. Data regarding the participants’ history of upper respiratory tract infection, any pre-existing illnesses, the type of surgery, and the anesthesia plan were recorded.

Intraoperatively, information related to the airway technique used, anesthetic agents administered, use of dexamethasone, size and type of airway material used, any difficulties encountered during airway instrumentation, number of attempts made, and any presence of blood on airway materials during intubation were documented.

Postoperatively, the duration of anesthesia and surgery, as well as complications such as laryngospasm, presence of blood on ETT or LMA at extubation, cough at extubation, stridor, hoarseness of voice, and vomiting after extubation, were recorded. After the child was discharged from the operating room and transferred to PACU, the presence or absence of sore throat, dysphagia, cough and hoarseness of voice were assessed using yes or no questions by the data collectors. The patient who developed at least one from postoperative sore throat, Postoperative hoarseness of voice or postoperative dysphagia were considered as pharyngeal compliant present.

### Study variables

Dependent variables:

- Postoperative pharyngeal complaints

Independent variables:

- Socio demographic characteristics: age, sex, weight and BMI
- Duration of anesthesia or surgery
- Type of airway material
- Inflation volume
- A number of attempts
- Blood on airway materials
- Cough at extubation
- Vomiting at extubation
- Use of dexamethasone
- Type of surgery

### Statistics

The statistical analyses were conducted using SPSS version 26 software packages. The normality of the continuous variable was assessed using the Shapiro-Wilk test. For non-normally distributed data, descriptive statistics such as medians and interquartile ranges were used to summarize the data. Since the data in this study did not follow a normal distribution, the results were presented as medians and interquartile ranges. The data were presented in frequency tables and graphs. To identify potential associated factors for postoperative pharyngeal complaint, both bivariate and multivariate analyses were performed using logistic regression. In the bivariate analysis, variables with a p-value < 0.25 were selected for further analysis in the multivariate analysis in order to identify strong predictors of postoperative pharyngeal complaint. A p-value less than 0.05 was considered statistically significant.

### Operational Definition

#### Postoperative pharyngeal compliant

If the patient has one or more from postoperative sore throat, dysphagia or postoperative hoarseness.

#### Postoperative sore throat

any discomfort or pain in throat during postoperative time

#### Dysphagia

any inability or difficulty to eat or swallow during postoperative time

#### Hoarseness

any voice change during postoperative time.

## RESULT

### Demographic and perioperative characteristics distribution

A total of 102 patients were included in this study, with 68 of them being male. The median weight of the patients was 24.5%, and the median age was 8 years. Most of the patients underwent endotracheal intubation ETT). The use of oral or nasal airway and face mask anesthesia were not reported. Cricoid pressure was applied in 21 patients (20.6%), and an introducer (stylet and bougie) was used in 24.5% of the included children.

All patients who underwent surgery under general anesthesia with laryngeal mask airway (LMA) used KY jelly for LMA cuff lubrication. Whereas lidocaine spray or injection was not applied in any of the cases. Propofol used for induction in almost half of the included patients, followed by a keto-fol. The predominantly used maintenance in the majority of patients was halothane (see table 1)

**Table 1.**
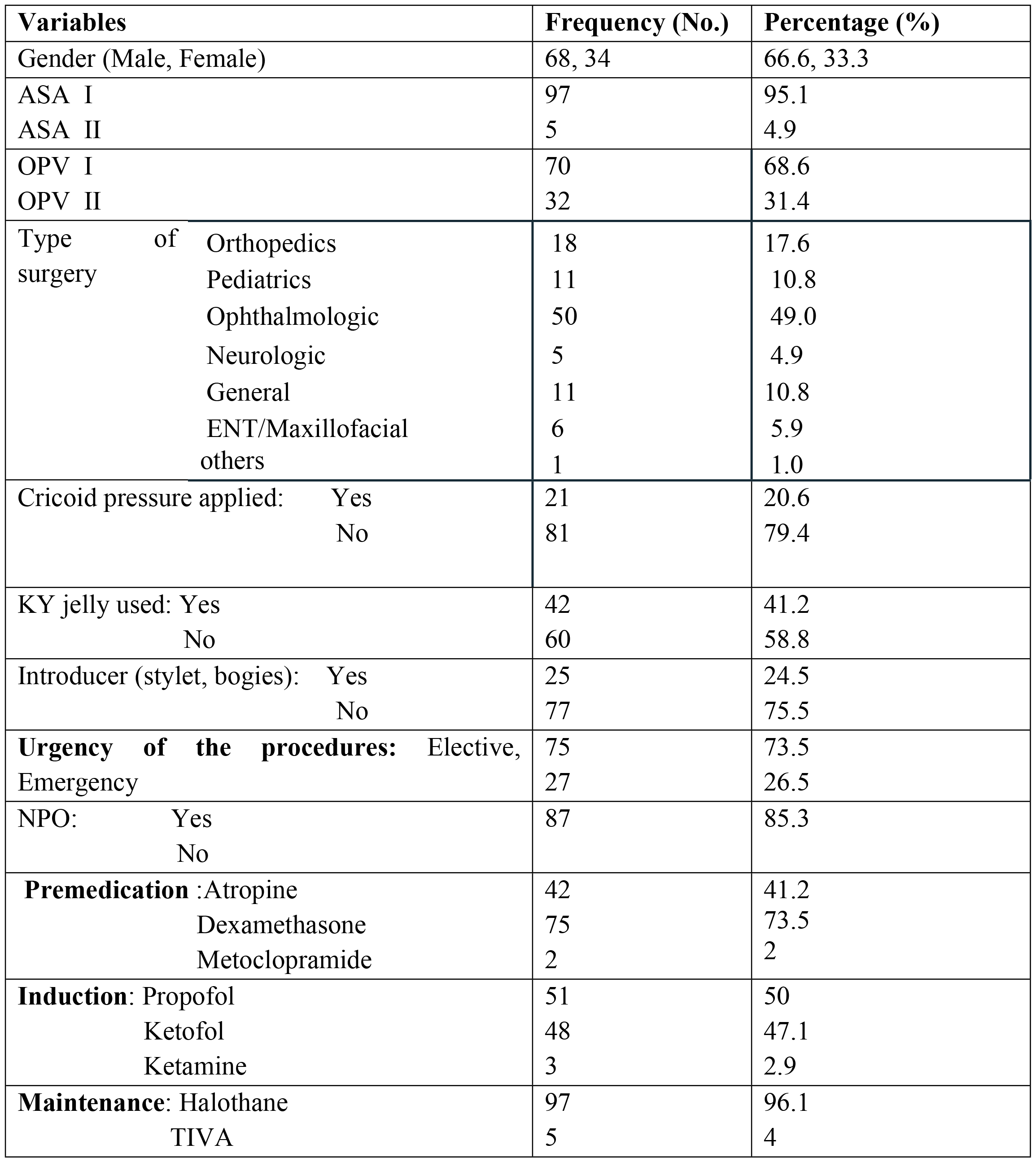
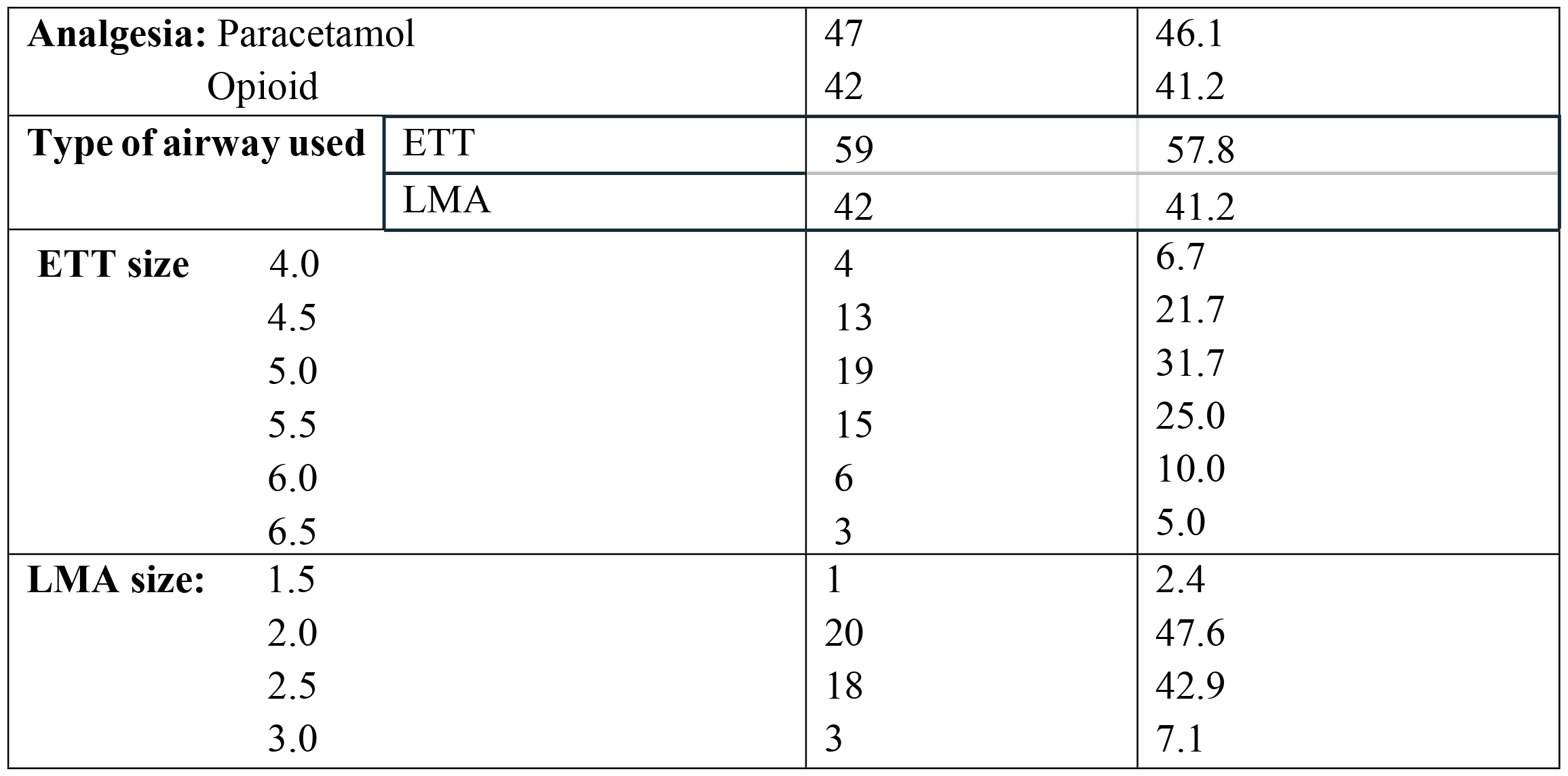
Socio-demographic and perioperative characteristics of the children operated under GA in HUCSH, Hawassa, Ethiopia from February-June 2022.

### Postoperative Outcome

The overall incidence of postoperative pharyngeal complaints is 32.4%. The specific incidence of common postoperative pharyngeal complaints were as follow; postoperative sore throat occurred in 27 patients (26.5%), cough 5.9%, Postoperative hoarseness 2.9% and dysphagia 4.9% were reported within the twenty-four hours postoperatively, regardless of severity scale. (fig.1)

**Figure 1.**
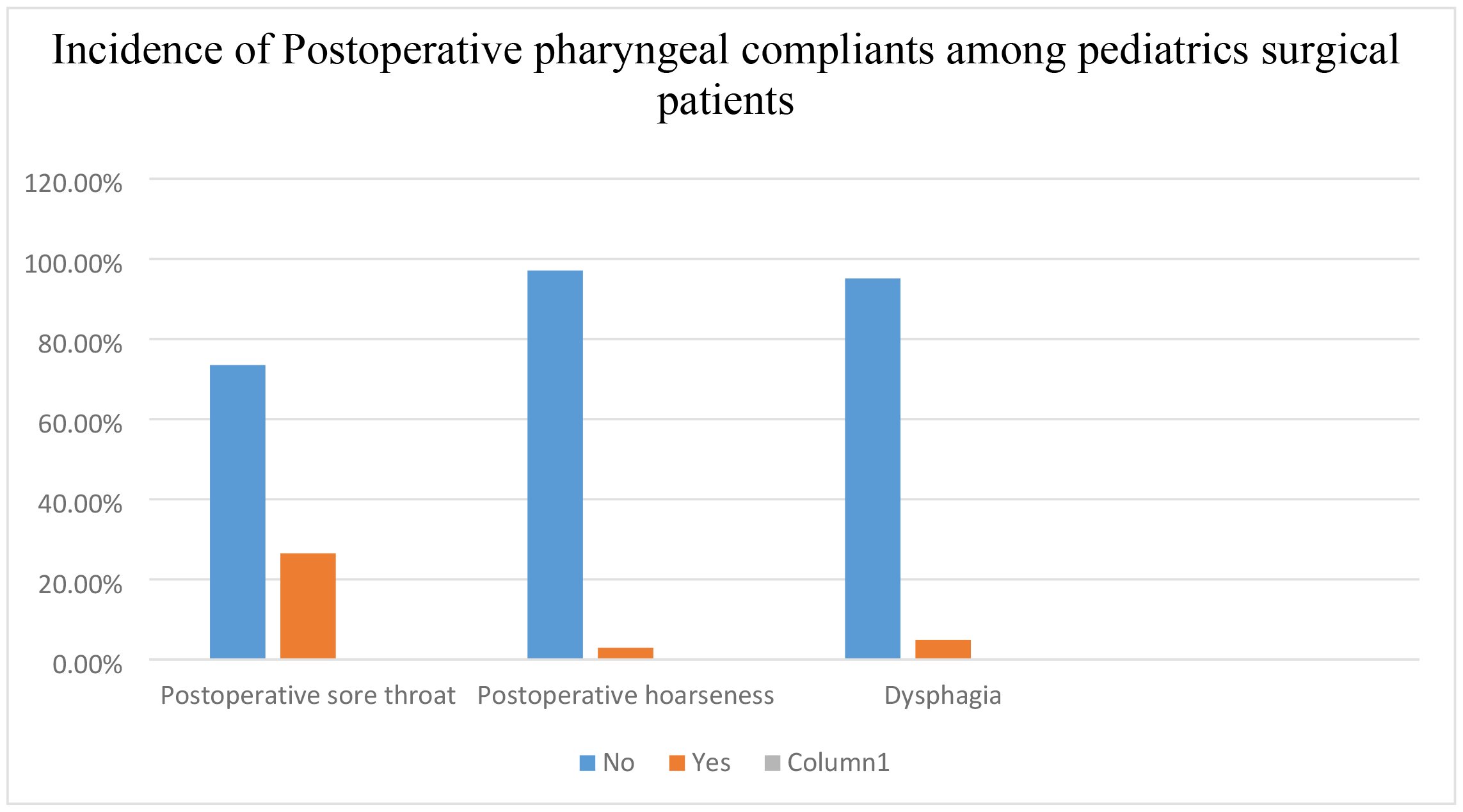
The incidence of postoperative pharyngeal compliant among pediatrics surgical patients that undergone surgery under general anesthesia at HUCSH, Hawassa, Ethiopia from February-June 2022.

### Predictors of postoperative Pharyngeal Complaints

Type of airway used, number of attempt, blood observed on laryngoscope during intubation and blood observed on ETT were associated with the incidence of postoperative pharyngeal complaints. (Table 2) However, certain independent variables such as gender, type of surgery, duration of intubation, vomiting on extubation, dexamethasone administration, and application of cricoid pressure were not found to be associated factors for postoperative pharyngeal complaints (p > 0.05). (Not included in table 2)

**Table 2.**
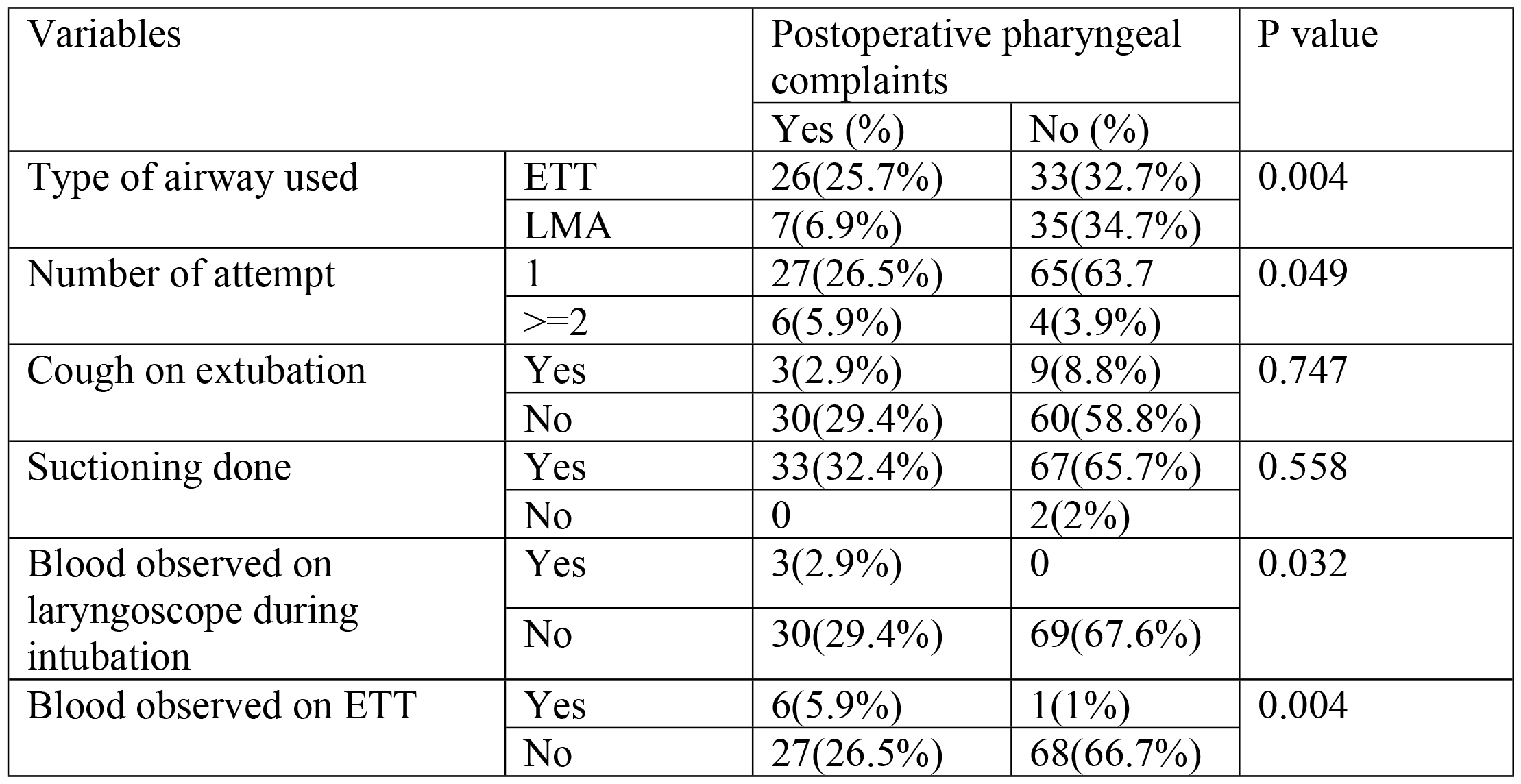
The associated factors of postoperative pharyngeal complaints among pediatrics surgical patients that undergone surgery under general anesthesia at HUCSH, Hawassa, Ethiopia from February-June 2022.

Multivariate analysis showed that endotracheal intubation was identified as the only statistically significant independent strong predictors of postoperative pharyngeal complaints with p-values of0.01 (AOR-3.846, 95% CI [1.385-10.682]).

## DISCUSSION

The result of this study revealed the overall incidence of postoperative pharyngeal complaints was 32.4%; whereas the prevalence of postoperative sore throat 26.5%, cough 5.9%, Postoperative hoarseness 2.9% and dysphagia 4.9% of participants.

In this study, the statistically significant independent predictors of postoperative pharyngeal complaints was endotracheal intubation (ETT) (p-values - 0.01, AOR-3.846, 95% CI [1.385-10.682]). The risk of developing postoperative pharyngeal complaint was found to be three times higher with endotracheal intubation compared to the use of a laryngeal mask airway (LMA).

The study conducted by Calder et al. reported an incidence of postoperative sore throat of 22.6% among pediatric patients who underwent surgery under general anesthesia using endotracheal intubation (15). This finding is consistent with the results of the present study, indicating a similar incidence rate of POST following ETT in pediatric surgical patients. Also our study shows ETT is strong predictors of postoperative pharyngeal complaints.

While there were no studies conducted within the same population as ours to compare these findings, the study done by Venugopal et al 2016 on adult population reported that the use of endotracheal intubation (ETT) was found to be associated with a higher incidence of voice problems and sore throat, whereas the use of a laryngeal mask airway (LMA) was associated with a higher incidence of dysphagia and odynophagia. (16). This finding is consistent with our study regarding high incidence of voice problem and sore throat in ETT, but our study did not show higher incidence of dysphagia and odynophagia in LMA.

In a previous study conducted by Calder et al. in the pediatric population with endotracheal tube (ETT) intubation, endotracheal tube cuff pressure was identified as a strong predictor of postoperative sore throat (p<0.0001) (15). However, in our cohort, cuff pressures for both ETT and laryngeal mask airway (LMA) were not monitored due to the absence of routine practices and equipment for cuff pressure measurement, making it difficult to assess the independent association of cuff pressure with postoperative pharyngeal compliant.

Instead of cuff pressure, we measured and analyzed the cuff inflation volume for LMA. However, when comparing the inflation volume to the maximum recommended by the manufacturer for each LMA size, we found no association between inflation volume and the occurrence of POST in this current study (p=0.328).

Another study by Wong et al., investigating the impact of LMA cuff pressure on the incidence of POST in children, demonstrated that hyperinflated LMAs resulted in increased airway morbidity compared to LMAs with lower inflation (10). The likely pathophysiology behind this finding is that hyperinflation of the LMA cuff leads to increased cuff pressure. If this pressure exceeds the pharyngeal wall perfusion pressure, it may result in pharyngeal ischemia, which can manifest as throat pain and dysphagia (17).

This study has some limitation including absence of monitoring cuff pressure which was strong predictor of postoperative pharyngeal complaint in previous studies, low sample size and heterogeneity of the study were another limitation of the study. Other airway techniques such as nasal intubation, oral and nasal airway were not found in our cohort because of the coincidence or practical matter.

## Conclusion

This study revealed the overall incidence of postoperative pharyngeal complaints was 32.4%. Endotracheal intubation was identified as the only independent predictors of postoperative pharyngeal complaints in children in this study.

However, we strongly recommend conducting large multicenter studies that assess the incidence, predictors, and impacts of postoperative complications in pediatric patients across all age groups, including infants, toddlers, and young children who may be unable to report their complaints.

## Data Availability

data will be available by contacting the corresponding author through email.

## Acknowledgements

I would like to express my appreciation for Hawassa university comprehensive specialized hospital for letting me to perform this study in this hospital and Dilla University for giving us the opportunity. We would also appreciate the willingness of data collectors, supervisors and study participants to participate in and play major role in this study.

## ABBREVIATIONS AND ACRONYMS

ASA: American society of Anesthesiologist
AOR: Adjusted odds ratios
CI: Confidence interval
ETT: Endotracheal tube
GA: General Anesthesia
HUCSH: Hawassa university comprehensive specialized hospital
OPV: Oropharyngeal view
PACU: Post anesthesia care unit
PH: Postoperative hoarseness
POST: Postoperative sore throat
LMA: Laryngeal mask airway
URTI: Upper respiratory tract infection

## Author’s contributions

The corresponding author(Adanech S Legasse) contributes in the study by conceptualization, study design, supervision, data analysis, and drafting the manuscript. Addisu Mossie, Aschalew Besha, and Nesra Mohammed have made a significant contribution in conceptualization, study design, analysis and editing the final draft. All Authors read and approved the final manuscript.

## Declaration of generative AI and AI-assisted technologies in the writing process

During the preparation of this work the authors used Chatgpt proofreading in order to paraphrase the article. After using this service, the authors reviewed and edited the content as needed and take full responsibility for the content of the publication.

## Notes

### Competing Interest Statement

The authors have declared no competing interest.

### Funding Statement

The author(s) received no specific funding for this work.

### Author Declarations

ethical clearance with reference number duchm/irb/055/2022 were secured from Dilla University institutional review board.

